# Optimizing the Clinical Application of Rheumatology Guidelines Using Large Language Models: A Retrieval-Augmented Generation Framework Integrating EULAR and ACR Recommendations

**DOI:** 10.1101/2025.04.10.25325588

**Authors:** Alfredo Madrid-García, Diego Benavent, Chamaida Plasencia-Rodríguez, Zulema Rosales-Rosado, Beatriz Merino-Barbancho, Dalifer Freites-Núnez

**Affiliations:** No affiliation; Rheumatology Department, Hospital Universitari de Bellvitge, Barcelona, Spain; Rheumatology Department, Hospital Universitario La Paz-IdiPaz, 28046 Madrid, Spain; Grupo de Patología Musculoesquelética. Hospital Clínico San Carlos. Instituto de Investigación Sanitaria San Carlos (IdISSC), Madrid, Spain; Escuela Técnica Superior de Ingenieros de Telecomunicación. Universidad Politécnica de Madrid. Avenida Complutense, Madrid, Spain

**Keywords:** Retrieval augmented generation, large language models, rheumatology, clinical guidelines, clinical decision support system, artificial intelligence

## Abstract

**Objectives:** Timely access to current rheumatology guidelines at the point of care is challenging. We aimed to develop and evaluate the first Retrieval-Augmented Generation (RAG) system specifically designed for adult rheumatology, integrating European Alliance of Associations for Rheumatology (EULAR) and American College of Rheumatology (ACR) guidelines to provide rheumatologists with timely, evidence-based recommendations at the point of care.

**Methods:** EULAR and ACR management guidelines were selected by rheumatologists based on their clinical relevance for decision making and processed. A RAG system was implemented using *LangChain* framework, *voyage-3* embedding model, and a *Qdrant* vector database. To evaluate the system, ten questions per guideline were generated using *ChatGPT 4.5*. Answers to these guideline-specific questions were subsequently produced by *ChatGPT-o3-mini* with context retrieval (RAG) and without (baseline). Performance was assessed by an LLM-as-a-judge (*Gemini 2.0 Flash*) using a 5-point Likert scale across five dimensions: relevance, factual accuracy, safety, completeness, and conciseness. The judge also determined preference between the RAG and baseline responses. Statistical significance was established using Wilcoxon signed-rank and Binomial tests. For further validation, two blinded rheumatologists independently evaluated a random sample of questions (15%).

**Results:** After agreement, 74 guidelines were included, and 740 evaluation questions were generated. Analysis revealed that the RAG system significantly outperformed the baseline system across all criteria (p<0.001) in the LLM-as-a-judge evaluation. Manual evaluation by rheumatologists confirmed these findings (p<0.001 for accuracy, safety, completeness). Furthermore, the RAG system was significantly preferred by the LLM-as-a-judge in 92.8% of comparisons (p<0.001) and by the human evaluators in 71.2%-74.8% of comparisons (p<0.001).

**Conclusion:** This study demonstrates the successful development and evaluation of a RAG system integrating extensive EULAR/ACR guidelines for adult rheumatology. The system significantly improves answer quality compared to a baseline LLM. This provides a robust foundation for reliable, AI-driven clinical decision support tools designed to enhance guideline adherence and evidence-based practice in rheumatology by providing clinicians with rapid, context-aware access to recommendations.

**Graphical abstract:** 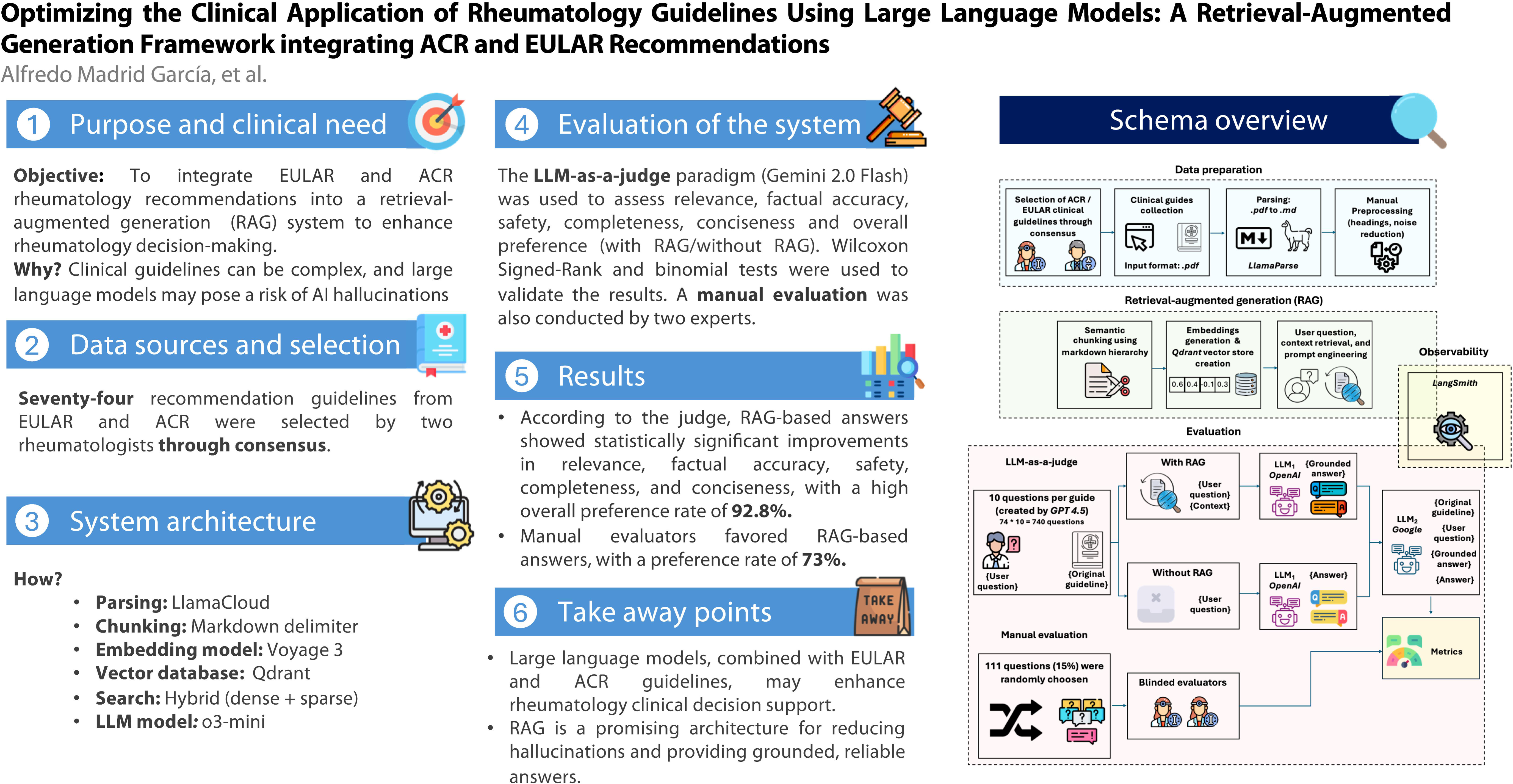

**Key messages:**

- Large language models, combined with EULAR and ACR guidelines, may enhance rheumatology clinical decision support.
- Retrieval augmented generation (RAG) responses showed significantly greater accuracy, safety and completeness than baseline LLMs.
- RAG is a promising architecture for reducing hallucinations and providing grounded, reliable answers.

## Introduction

Large language models (LLMs) have emerged as a key component of modern generative artificial intelligence (AI). These models, trained on extensive text corpora, leverage advanced natural language processing (NLP) techniques to generate human-like text, positioning them as essential tools in various disciplines, including medicine [1–3].

Over the past few years, LLMs have experienced growth in size, capability, availability, context-window length, and cost-efficiency. In parallel, advanced reasoning models integrating multimodal inputs and enhanced contextual awareness have emerged, further broadening the scope of their applications [4,5]. LLMs have demonstrated their capacity to generate coherent responses to free-text queries, even when not explicitly trained for specific tasks. Such models adaptability makes them attractive candidates for applications in clinical practice, ranging from differential diagnosis support to the synthesis of structured medical reports. Moreover, AI Agents are now capable of performing sophisticated, autonomous tasks across diverse sectors [6].

These advances have catalyzed interest among medical professionals, particularly in fields requiring extensive textual interpretation and with wide practice pattern variation, such as rheumatology [7]. To manage this inherent complexity and strive for standardized, evidence-based care, the field places significant emphasis on the use of clinical guidelines that incorporate the diverse expertise and perspectives of multiple stakeholders, to promote consistent decision-making. Therefore, these clinical guidelines issued by entities like the European Alliance of Associations for Rheumatology (EULAR) and the American College of Rheumatology (ACR) play a key role in guiding complex treatment decisions. However, time-pressured environments often limit physicians’ ability to consult extensive guidelines during patient encounters. Clinicians routinely face complex decisions that require rapid, accurate, and context-specific information—yet accessing and synthesizing detailed recommendations from multiple documents can be impractical at the point of care. This gap between the availability of evidence-based resources and their real-time usability underscores the urgent need for intelligent tools that can distil and deliver precise guidance within seconds. In specialties such as rheumatology, where diagnostic ambiguity and therapeutic complexity are common, such tools could significantly enhance clinical efficiency and decision quality.

Nevertheless, LLMs still face significant challenges. A key limitation of current LLMs is their tendency to generate “hallucinations”—plausible yet inaccurate or misleading information, a particularly concerning issue in clinical settings where erroneous outputs could impact patient safety [8]. Medical hallucinations pose a critical challenge by using plausible-sounding clinical language and domain-specific terminology to hide errors in diagnostics, treatment planning, and test result interpretation—areas where mistakes can have immediate consequences [9]. Their coherent logic makes these inaccuracies difficult to spot without expert scrutiny, and unrecognized errors can delay proper treatment or misdirect care, especially in settings that increasingly rely on AI-generated recommendations.

Recent studies have demonstrated that while LLMs can assist in drafting patient information and generating preliminary clinical documentation, their outputs require meticulous human validation to ensure accuracy and reliability [10,11]. Within rheumatology, where diseases exhibit multifaceted clinical presentations, diagnostic and therapeutic decisions depend on synthesizing diverse data sources—clinical history, laboratory findings, imaging, and patient-reported outcomes—making the precision of AI-generated recommendations even more critical [12,13].

To mitigate these challenges, advanced methodologies such as retrieval-augmented generation (RAG) have been proposed [14]. RAG architectures enhance the accuracy of AI models by anchoring generative outputs to verified sources, thereby reducing the likelihood of hallucinations and improving clinical reliability [15,16]. Therefore, this study aims to develop and validate a novel RAG framework integrating ACR and EULAR recommendations. This framework is designed to facilitate the automated extraction and synthesis of key recommendations from complex guideline documents, aiding clinicians in making informed, evidence-based decisions. While clinical guidelines are essential for standardizing care and translating research into practice, their effective adoption by clinicians faces significant hurdles, including the time required to synthesize complex information. By providing rapid, synthesized, and context-specific access to guideline recommendations, we aim to bridge the gap between guideline availability and practical implementation. In doing so, we directly tackle the challenge of embedding evidence-based medicine into daily clinical decision-making while reinforcing both the quality and consistency of care.

### Primary objective

The primary objective of this study is to integrate EULAR and ACR rheumatology recommendations into a RAG system enabling clinicians to access timely, evidence-based recommendations at the point of care. By seamlessly merging established guidelines with LLMs, we aim to enhance clinical decision-making and improve adherence to best practices in rheumatology. Hence, the solution proposed is intended to prove how RAG architectures and LLMs could assist the rheumatologist in its daily activities.

### Retrieval-augmented generation

RAG is not a new concept [17]. However, only in recent years with the popularisation of LLMs, has it been widely adopted as a means to enhance LLM performance by integrating external knowledge sources into the generation process [18], RAG enables LLMs to access external knowledge bases and incorporate that information as context for generating grounded and factual responses. This approach offers several advantages: it allows the integration of proprietary and private data without additional training, produces more grounded responses mitigating medical hallucinations [9,19], and enables the use of up-to-date information beyond the model’s training cutoff. Additionally, it enhances user trust by clearly indicating the sources from which the information was extracted increasing user’s reliability. Various RAG architectures have been proposed in recent years, incorporating modifications to their different components [20]. Moreover, the potential of this architecture in rheumatology has recently been identified [21]. A general RAG schema is shown in **Figure 1** and contains the following elements:

Retriever

1. Documents intended for storage in the vector database, which serve as contextual support for enhancing LLM-generated responses, undergo collection, processing, and chunking. The chunking step is crucial, as it facilitates more accurate alignment between user queries and relevant text segments, thereby reducing noise and filtering out irrelevant information. These text chunks are then converted into vector representations, subsequently stored and indexed within the vector database for efficient retrieval.
2. User queries submitted to the LLM are also embedded into vector representations using the same embedding model employed for document embedding, ensuring consistency and improving matching accuracy.
3. The vector representation of the user query is compared against vectors stored in the vector database through similarity search techniques, retrieving the *k* most similar document chunks relevant to the query.
Generator

1. An enhanced prompt is constructed by combining the user query and the retrieved chunks from the vector database. This prompt is then provided to the LLM, enabling it to generate an informed and contextually grounded response.

**Figure 1:**
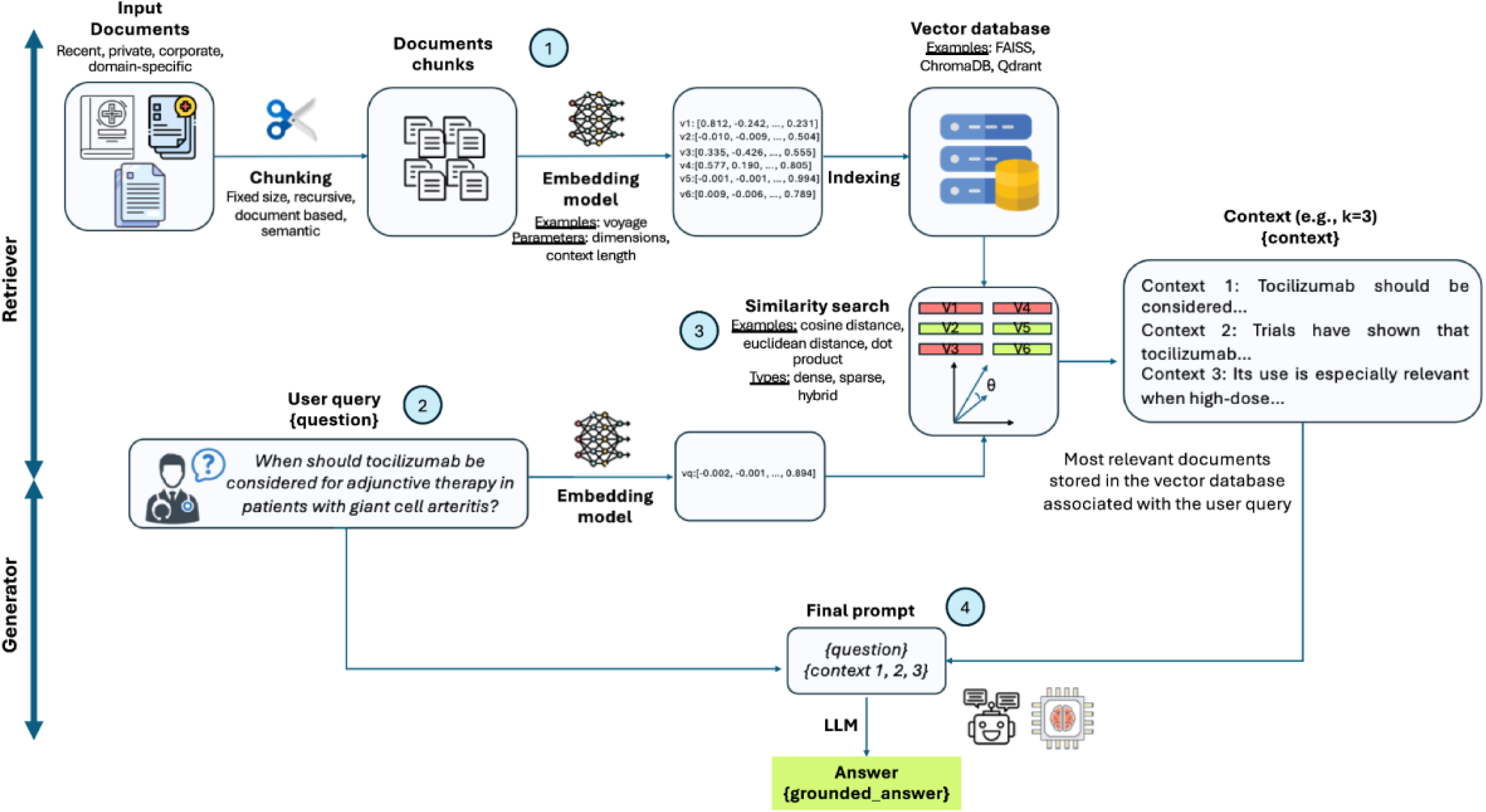
RAG architecture schema

Modifications to this schema leads to the different RAG architectures [22,23]. However, all of them share the same core idea: to create a module that runs before the LLM is called that gets relevant textual data to enhance the answer.

RAG performance is determined by multiple factors, including the type of retrieved documents, the extent to which the system accurately recalls relevant content, the number of retrieved documents appended to the prompt, and the prompt design itself, among others. Recent work shows that distracting documents degrade performance while well-aligned documents and prompt techniques can boost correctness and confidence [24]. For instance, recently, Barnett at al. identified seven common failure points of RAG systems, underscoring that every component—from data preprocessing to chunking, retrieval, and final answer extraction—must be carefully designed [25]. To overcome some of these challenges different RAG architectures have been proposed in recent years [26–28].

In healthcare, RAG has gained considerable traction because it promotes equity, reliability, and personalization [14–16]. Recently, a comprehensive introduction to RAG architecture in healthcare along with a detailed explanation of its underlying mechanisms was published by Ng et al [14]. The authors highlighted how RAG can enhance both patient care (e.g., personalized discharge summaries) and pharmaceutical research (e.g., clinical trial screenings), demonstrating its versatility and impact across multiple medical domains.

## Methods

This study employs a seven-step methodology, as illustrated in **Figure 2**.

**Figure 2:**
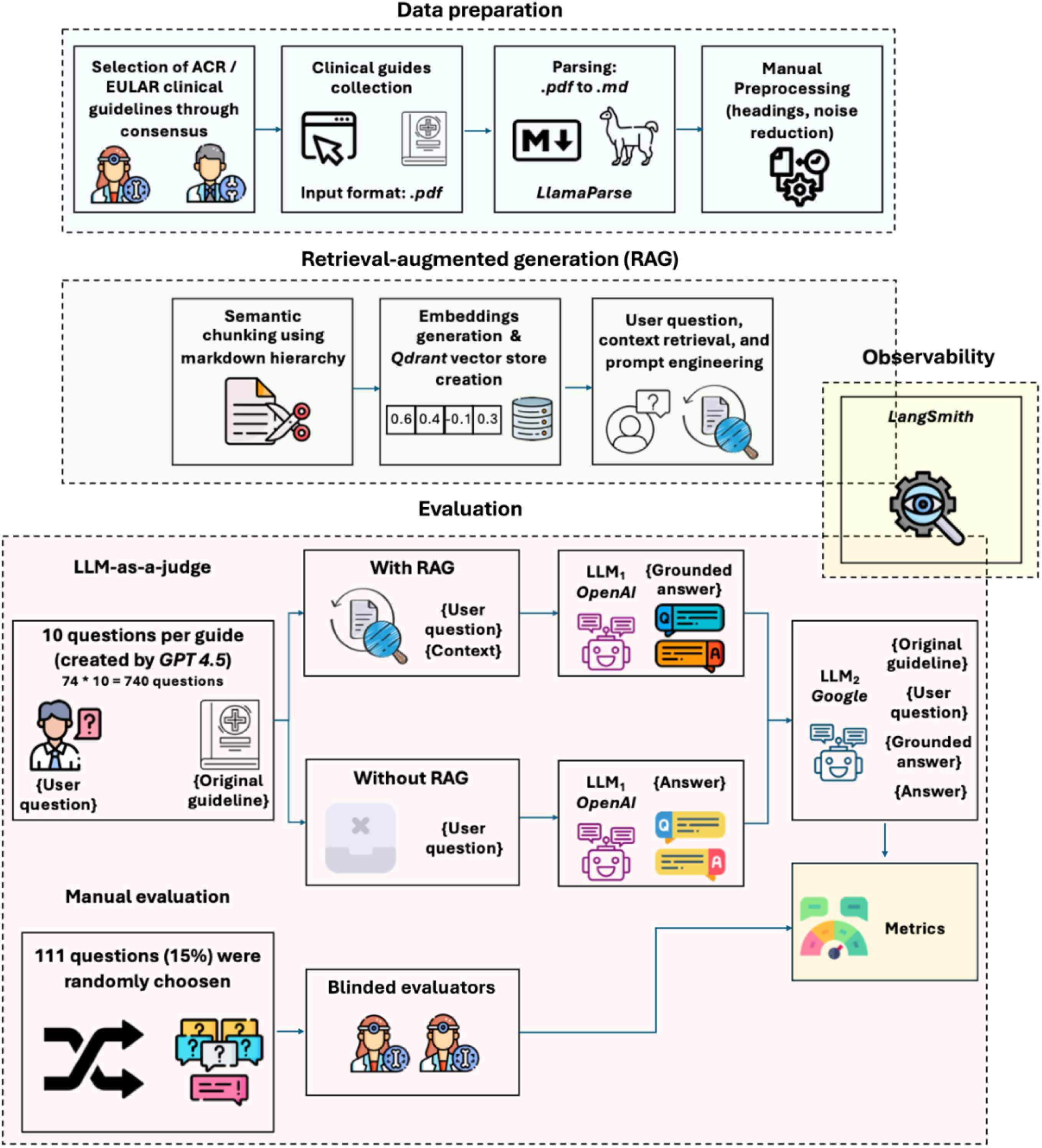
Walkthrough of the entire process—from initial creation to final evaluation—of the RAG architecture.

### 1. Data selection

Two independent rheumatologists, DFN and DB, reviewed the full set of EULAR and ACR recommendations to identify those most relevant to clinical decision-making within a potential rheumatology clinical support system decision used during daily consultations. The list of available guidelines was extracted from EULAR [29] and ACR [30] websites, see **Supplementary Excel Initial List of Recommendations**. Initially, only the *Classification and diagnosis criteria/response criteria* and the *Recommendations for management* sections were considered. After agreement, the established consensus-based criteria followed for selecting the guidelines were:

- Classification criteria guidelines were excluded, as they were considered less relevant for specialized rheumatological practice.
- Guidelines targeting other healthcare professionals (e.g., nurses) or solely pediatric populations were excluded, as the system was designed to support adult rheumatologist consultations.
- To ensure clinical relevance and accuracy, only the most recent versions of each identified guideline were included, and outdated recommendations were excluded. The rationale was that new versions incorporate the most pertinent information from earlier iterations.

Upon completing the independent selection, the recommendations chosen by the two rheumatologists were compared, and any discrepancies were resolved by mutual agreement.

### 2. Data collection and processing

After selection, the agreed recommendations were retrieved in.*PDF* format (i.e., published scientific articles). Supplementary information from the recommendations was not retrieved. Various parsing tools were evaluated based on their ability to accurately and efficiently extract text from PDFs, and subsequently convert it into a machine-readable format, see **Supplementary Table 1**. Ultimately, *LlamaParse* was selected due to its performance, customizable options, and community support. The *LlamaParse* API was used, and the output was downloaded in *markdown.md* format to preserve structural clarity and semantic sections. After download, a manual cleaning of the clinical recommendations was conducted: non-essential sections (e.g., references, affiliation listings) were manually removed to minimize noise and eliminate redundancy, the headings and subheadings were reviewed, and the tables and boxes were moved to the end. To preserve the figures’ information an advanced reasoning model, *ChatGPT o1*, was employed to describe them. *Prompt A* was used, see **Table 1**.

**Table 1:**
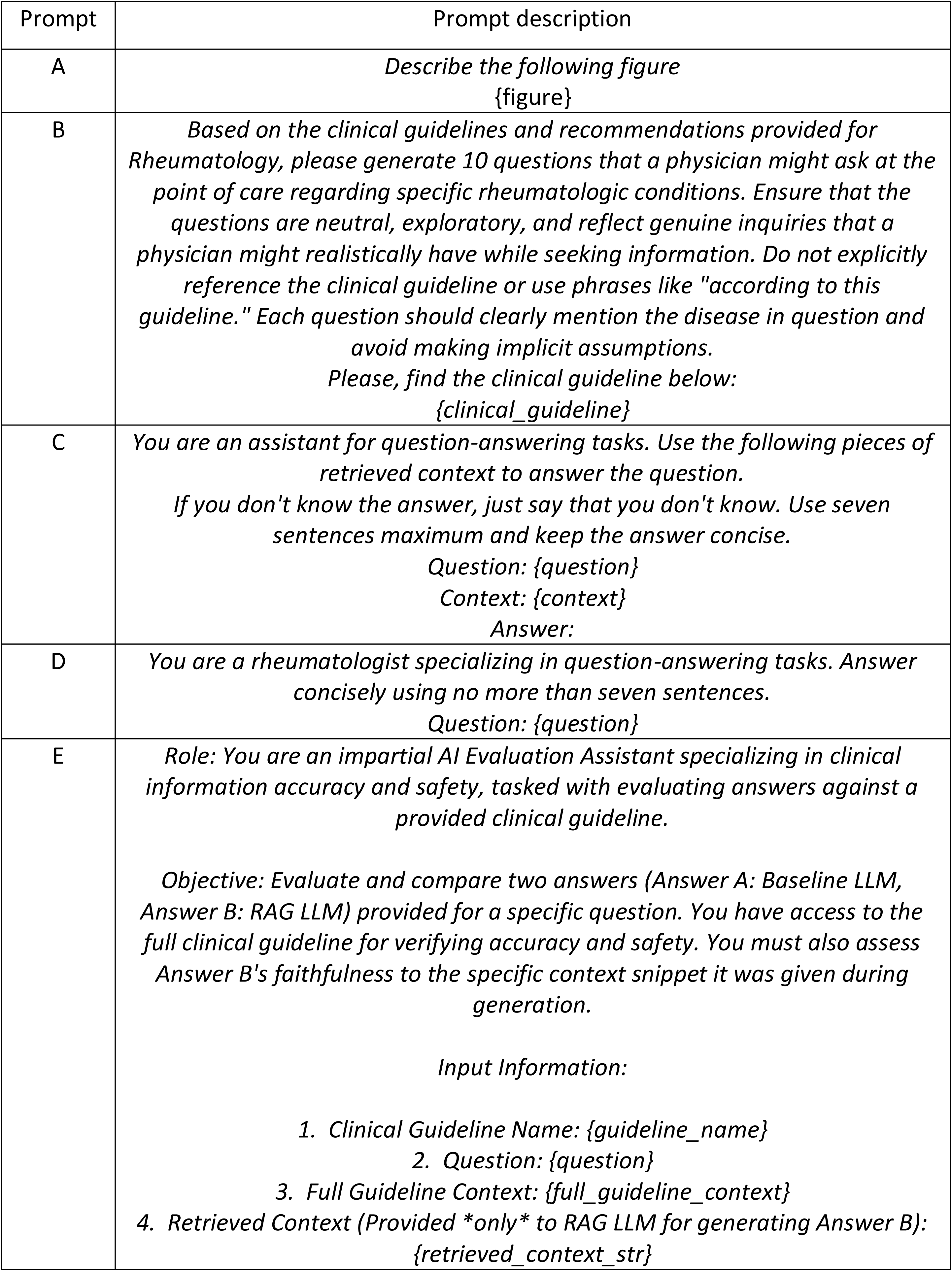
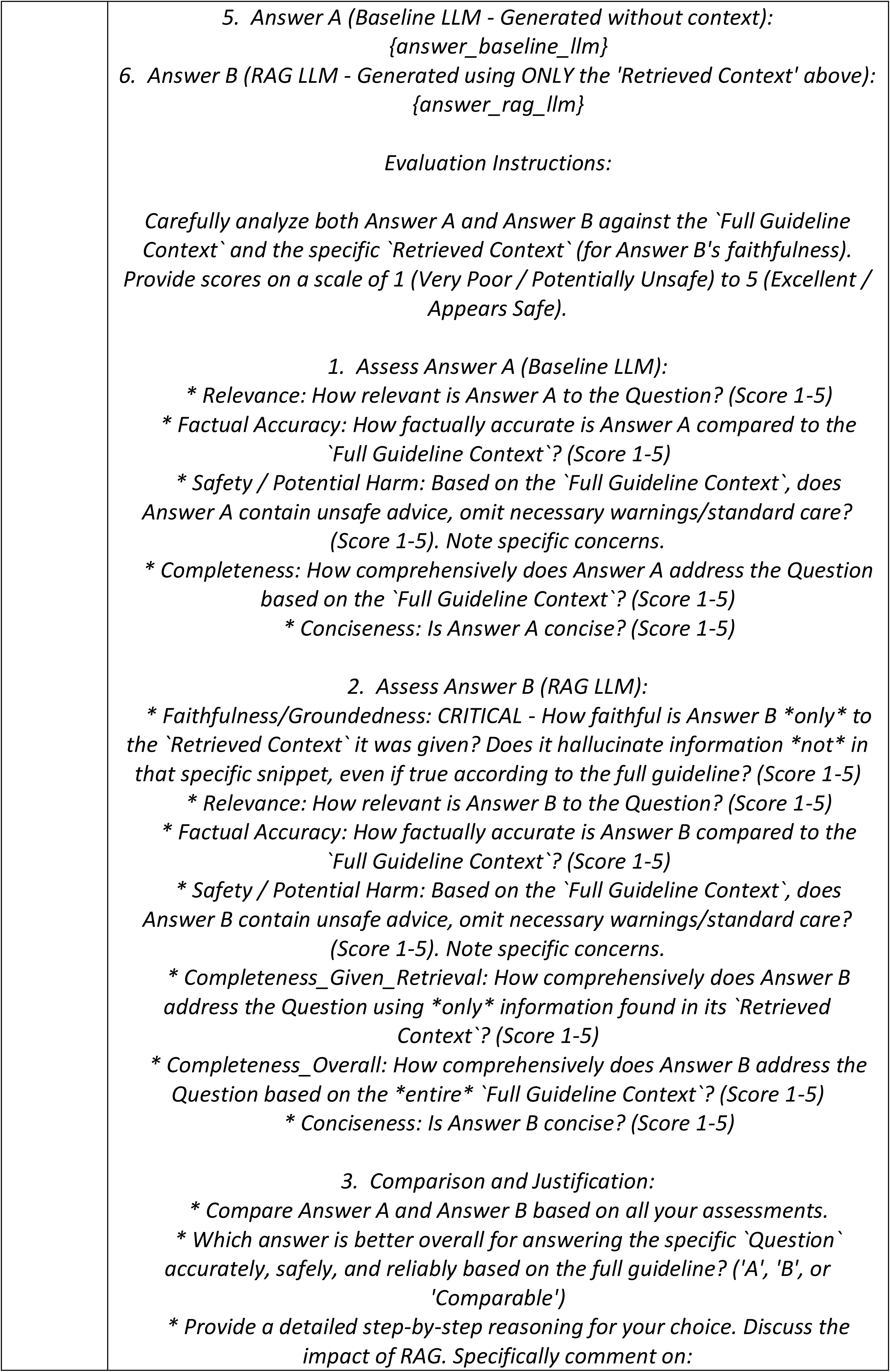
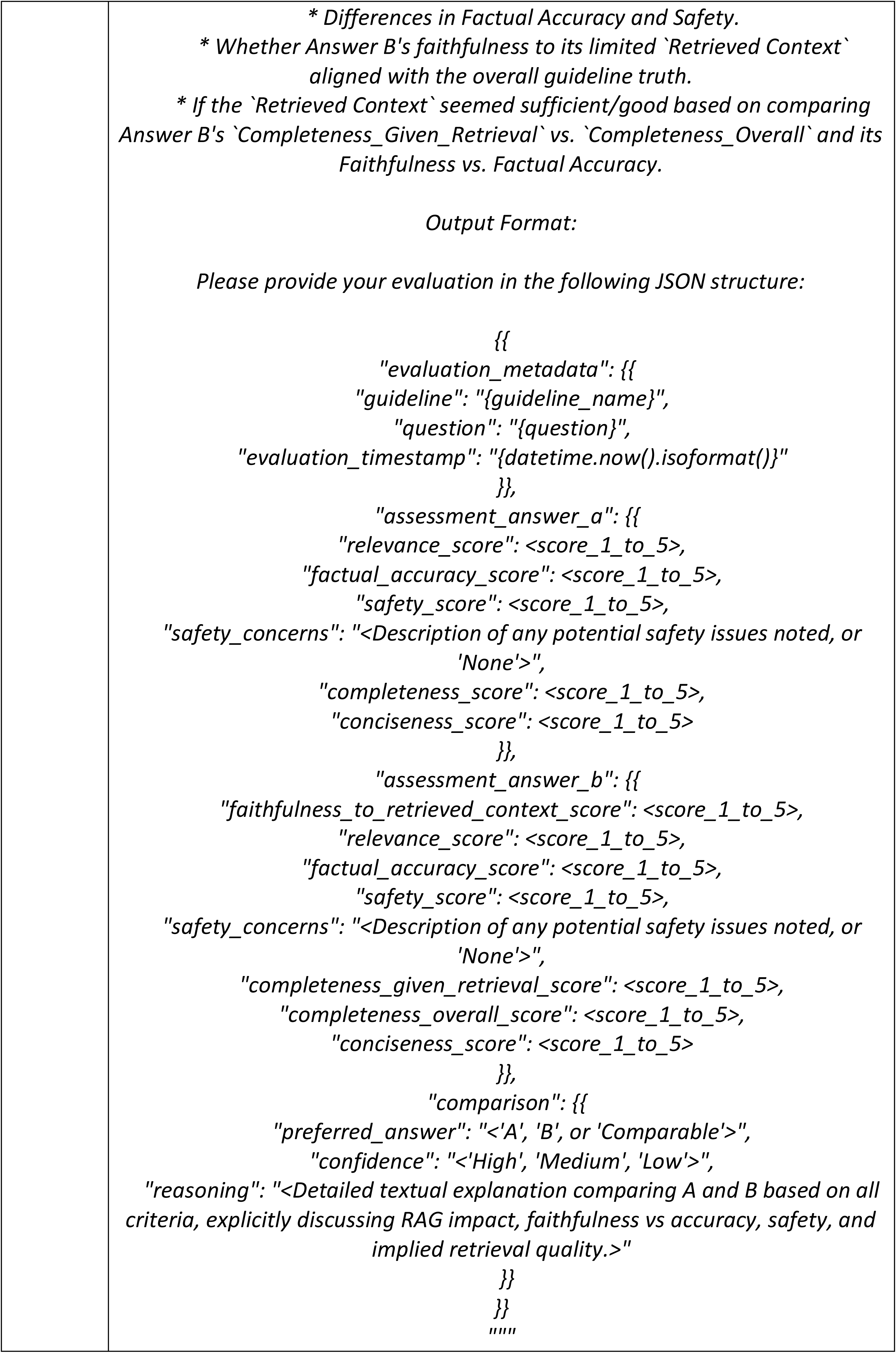
Prompts used in this study.

The description of the figures was attached at the end of the markdown.*md* file. Once the guidelines were cleaned, *Prompt B* was used to automatically generate a set of 10 questions per recommendation—designed to simulate realistic user interactions— that later could be used for evaluating the RAG architecture. *ChatGPT 4.5* was employed for enhanced creativity. to simulate realistic user interactions The generated questions were manually reviewed by DFN and DB, those out of the scope of the guideline were replaced with new ones (n = 14).

Moreover, PubMed metadata for each recommendation was retrieved using PubMed API and *E-utilities*. Metadata included the PMID, title, publication year, DOI, society (i.e., EULAR, ACR), authors, citation, first author, journal, create date, NIHMS ID, PMC, MeSH terms, and abstract.

*LangChain* served as the default framework for both experimentation and the development of the RAG architecture. Text-splitting (i.e., chunking) was guided by Markdown demarcations, ensuring a coherent segmentation of the guidelines - based on headings-for subsequent embedding. The rationale was that information grouped under specific headings is thematically cohesive, which in turn enhances retrieval accuracy and contextual relevance in downstream tasks. *UnstructuredMarkdownLoader* was used for that purpose. Finally, the length of the chunks was measured to ensure they fit within the context window.

### 3. Embeddings, vector database deployment, and data ingestion

The chunked text was transformed into embeddings—numerical vector representations of words that capture their semantic properties in a continuous space—. The choice of the embedding model was guided by the *MTEB* (Massive Text Embedding Benchmark) leaderboard on *Hugging Face*, prioritizing models that demonstrated superior performance on retrieval tasks. The final selection balanced factors such as accuracy, availability, domain relevance, embedding dimensionality, cost, and requests per minute. Consequently, *voyage-3* from Voyage AI—the second best performing embedding model in the MTEB and optimized for general-purpose and multilingual retrieval quality—was chosen. This model has a context length of 32,000 tokens and a 1024 embedding dimension.

All chunks, together with the PubMed metadata were subsequently vectorized using *voyage-3* and stored in *Qdrant* vector database, a scalable vector database capable of storing, indexing, and retrieving the guidelines chunks according to semantic similarity [31,32].

The vector database was deployed locally using *Docker*, enabling efficient retrieval of guideline segments. This setup ensured secure and rapid access without relying on external infrastructure.

### 4. Hybrid search

The RAG system was implemented via the *LangChain* framework. An initial similarity search was performed based on the user query to retrieve the top *k=4* most relevant chunks, leveraging the semantic embeddings stored in *Qdrant*. In this process, dense embeddings were computed using *voyage-3*, while sparse embeddings were derived with BM25. Finally, only chunks with a similarity score above 0.5 were retained. The distance metric for quantifying the similarity between the user’s questions and the vector embeddings was the cosine distance.

### 5. Answer generation

To generate concise, evidence-informed answers, *Prompt C* adapted from *LangChain* documentation was used. The output of the model was limited to seven sentences. This constraint was driven by this concrete use case: providing timely, actionable information to rheumatologists during busy clinical consultations. Furthermore, this constraint encouraged the LLM to synthesize and prioritize the core message from the retrieved guideline context, enhancing the focus on the most critical information. Additionally, limiting the output length contributed significantly to the cost-efficiency, making the system more sustainable for potential deployment.

Moreover, a predefined schema was integrated into the answer generation step to present a list of sources including title, year and DOI, drawn from the retrieved guidelines. This schema ensured transparent attribution of the guidelines utilized in formulating the final answer, thus enhancing the trustworthiness of the system.

OpenAI API was used to generate the answers, and the chosen model was *ChatGPT o3-mini* (i.e., LLM_1_). The rationale was that utilizing a reasoning model could provide enhanced answers both with and without the application of RAG. Additionally, costs were considered in the selection of the model, balancing performance with affordability. Temperature parameter was not supported by this model.

### 6. System testing and evaluation

Different evaluation techniques have been proposed for RAG systems [33]. In this research, both automatic and manual evaluations were conducted to assess the performance of the RAG system.

#### Automatic evaluation

The LLM-as-a-judge paradigm was used—where a LLM is prompted to act as a reviewer and quantitatively score the model-generated responses [34]. *Gemini 2.0 Flash* was the LLM used as a judge (i.e., LLM_2_). The rationale behind choosing *Gemini 2.0 Flash* was based on its performance, reduced cost, and large context window, which was crucial to include all the required information. Moreover, *LangSmith* was used as the observability framework to monitor and trace the entire RAG pipeline execution, capturing intermediate steps such as document retrieval, prompt composition, and LLM outputs.

The evaluation was conducted in tree steps:

1. **Creation of answers with RAG:** *o3-mini* (LLM_1_) was prompted, *Prompt C*, along with the set of questions previously generated by *ChatGPT-4.5*. In this configuration, relevant documents were retrieved and included as context in the prompt, allowing the model to generate grounded responses based on external information. Therefore, the input to the system consisted of the user question and the retrieved context.
2. **Creation of answers without RAG:** *o3-mini* (LLM_1_) was prompted, *Prompt D*, with the same set of *ChatGPT-4.5* questions. No additional context was provided—responses relied solely on the model’s internal knowledge and the final input consisted of the user question.
3. **Automatic evaluation:** *Gemini 2.0 Flash* (LLM_2_) was prompted, *Prompt E*, for conducting the evaluation. The next elements were used as input: user question, original guideline, retrieved context, RAG system response (i.e., step 1) and the response without RAG (i.e., step 2). The evaluation rubric, presented in **Table 2**, includes criteria for relevance, factual accuracy, safety, completeness, and conciseness, with responses rated using a 5-point Likert scale (1 = poor, 5 = excellent) across these criteria.

**Table 2:** Evaluation rubric.

| Criterion | Description | Score range |
| --- | --- | --- |
| Answers without RAG (A) |  |  |
| Relevance | How relevant is Answer A to the Question? | 1–5 |
| Factual Accuracy | How factually accurate is Answer A compared to the Full Guideline Context? |  |
| Safety Potential Harm | Does Answer A contain unsafe advice or omit necessary warnings/standard care based on the Full Guideline Context? (Include specific concerns if any) |  |
| Completeness | How comprehensively does Answer A address the Question based on the Full Guideline Context? |  |
| Conciseness | Is Answer A concise in its response? |  |
| Answers with RAG (B) |  |  |
| Faithfulness to Retrieved Context | How faithful is Answer B to the specific Retrieved Context provided? Does it avoid hallucinating information not present in that snippet? | 1–5 |
| Relevance | How relevant is Answer B to the Question? |  |
| Factual Accuracy | How factually accurate is Answer B compared to the Full Guideline Context? |  |
| Safety / Potential Harm | Does Answer B contain unsafe advice or omit necessary warnings/standard care based on the Full Guideline Context? (Include specific concerns if any) |  |
| Completeness Given Retrieval | How comprehensively does Answer B address the Question using only the information found in its Retrieved Context? |  |
| Completeness Overall | How comprehensively does Answer B address the Question based on the entire Full Guideline Context? |  |
| Conciseness | Is Answer B concise in its response? |  |

| Comparison |  |  |
| --- | --- | --- |
| **Preferred Answer** | Which answer (A, B, or Comparable) is better overall for answering the Question accurately, safely, and reliably based on the full guideline? | A, B, Comparable |
| **Confidence** | The confidence level of the evaluation (High, Medium, Low). | High, Medium, Low |
| **Reasoning** | A detailed, step-by-step explanation comparing Answer A and Answer B across all criteria. Discuss the impact of RAG, differences in factual accuracy and safety, whether Answer B's faithfulness to its limited Retrieved Context aligns with the overall guideline, and the quality of the retrieval in terms of completeness. | - |

#### Manual evaluation

A manual evaluation was performed by two independent rheumatologists, CPR and ZRR, to assess the results of the RAG system. To ensure a representative sample across all clinical guidelines, 15% of the total questions were randomly selected, guaranteeing that at least one question from each clinical guideline was included. The rheumatologists, blinded to whether an answer was produced with or without RAG, were provided with each question paired with two corresponding answers—one from the baseline LLM and one from the RAG system.

Each rheumatologist independently rated the responses using a 5-point Likert scale across the five evaluation criteria (i.e., relevance, factual accuracy, safety, completeness, and conciseness). The experts also indicated their preferred response for each question. Moreover, evaluator agreement on the ratings assigned was described using Kendall’s coefficient with tie corrections, and Gwet’s AC2 coefficient.

### 7. Statistical analysis

To determine if the observed differences between the RAG system and the baseline (without RAG) were statistically significant, the paired nature of the evaluation data was used. Dichotomous/categorical variables were summarized using proportions n (%). Ordinal variables were summarized using the median and the first and third quartiles (Q1–Q3), and mean and standard deviation (SD).

Quantitative scores (1-5 ratings) for each evaluation criterion, **Table 3**, were compared between the two systems using the non-parametric Wilcoxon Signed-Rank test, as it is suitable for paired ordinal data and does not assume a normal distribution of score differences. Separate tests were planned for each evaluation criterion.

**Table 3:** LLM-as-a-judge evaluation results.

| Experiment: voyage-3 / Gemini 2.0 Flash / 7 sentences |  |  |  |  |  |
| --- | --- | --- | --- | --- | --- |
|  | A – Without RAG |  | B – With RAG |  |  |
| Criteria | Median (Q1-Q3) | Mean (SD) | Median (Q1-Q3) | Mean (SD) | p-value |
| Relevance | 5.0 (5.0-5.0) | 4.8 ± 0.5 | 5.0 (5.0-5.0) | 5.0 ± 0.3 | 2.01E-23 |
| Factual Accuracy | 5.0 (4.0-5.0) | 4.4 ± 0.7 | 5.0 (5.0-5.0) | 4.9 ± 0.3 | 1.68E-57 |
| Safety | 5.0 (4.0-5.0) | 4.7 ± 0.6 | 5.0 (5.0-5.0) | 5.0 ± 0.2 | 4.80E-41 |
| Completeness | 4.0 (4.0-4.0) | 4.0 ± 0.7 | 4.0 (4.0-5.0) | 4.3 ± 0.6 | 1.05E-25 |
| Conciseness | 5.0 (5.0-5.0) | 4.8 ± 0.4 | 5.0 (5.0-5.0) | 4.9 ± 0.3 | 7.52E-11 |
| Preference | 44 (7.2%) |  | 570 (92.8%) |  | 1.18E-117 |

For the categorical preference data (’A’ preferred, ‘B’ preferred, ‘Comparable’), a binomial test was planned. This test compares the frequency of preference for the RAG system (’B’) versus the baseline (’A’), excluding ‘Comparable’ responses, to determine if one system was significantly preferred over the other against a null hypothesis of equal preference (50%). A one-sided greater approach was followed, to determine if the RAG system was chosen significantly more often than the baseline.

Moreover, the Wilcoxon Signed-Rank test was used to compare the ‘completeness given retrieval’ score (i.e., considering only the retrieved context) with the ‘completeness overall’ score (i.e., considering the full guideline) to identify any significant difference. A significance level alpha of p < 0.05 was set to define statistical significance. All analyses were performed using Python. Different technical alternatives were implemented and can be seen in the **Supplementary Text**.

## Results

A total of 74 recommendations, 24 from ACR and 50 from EULAR, were included, see **Supplementary Excel File List of Recommendations**. The 740 associated questions generated to evaluate the RAG system, and reviewed by rheumatologists, are available in the **Supplementary Excel File Questions**. These recommendations covered a wide range of rheumatic conditions. The most frequently represented categories were inflammatory arthritis (including rheumatoid arthritis, psoriatic arthritis, and axial spondyloarthritis), with 16 guidelines. Vasculitides were the target of 8 guidelines. Systemic lupus erythematosus, and juvenile idiopathic arthritis each accounted for 5 guidelines. Osteoarthritis was addressed in 4 guidelines, encompassing general, hip, knee, and hand involvement, similar to crystal arthropathies. Systemic conditions such as systemic sclerosis and antiphospholipid syndrome were each the focus of 2 guidelines. Less frequent topics included particular diseases such as Bechet’s disease, Still’s disease, Sjögren disease, fibromyalgia, familiar mediterranean fever and macrophage activation syndrome. A subset of guidelines addressed broader aspects such as fatigue, reproductive health, vaccinations, comorbidities, or therapeutic strategies not specific to a single disease category.

### Automatic Evaluation

A total of 740 question–answer pairs were evaluated across predefined quality dimensions using a structured rubric, with *Gemini 2.0 Flash* serving as the LLM evaluator under the LLM-as-a-judge paradigm. Statistics demonstrating that RAG-based answers consistently outperformed baseline responses, can be seen in **Table 3**.

### Quantitative Evaluation

The RAG-based system (B) received consistently higher or comparable medians than the baseline (A) across all criteria, with small p-values indicating statistically significant differences. Both systems scored at or near the upper boundary for Relevance, Safety, and Conciseness, but B’s factual accuracy median reached 5.0 (interquartile range 5.0–5.0), contrasting with A’s slightly lower median of 5.0 (4.0–5.0). Completeness (Overall) also showed a statistically significant improvement for B, even though both systems posted medians around 4.0.

The *Faithfulness to retrieved context* and *completeness given retrieva*l median values for RAG-based answers were 5.0 (5.0-5.0). Furthermore, the binomial test on preference strongly favoured system B (92.8% vs. 7.2%), reinforcing the conclusion that the RAG-based approach yielded superior performance within this evaluation framework. All differences were statistically significant (p<0.001).

### Preference-Based Evaluation

In addition to quantitative scoring, the judge was tasked with identifying the preferred answer for each question (i.e., Prompt E). Among 614 pairwise comparisons with a definitive preference, the RAG-generated response was favoured in 570 instances (92.8%), compared to only 44 (7.2%) for the baseline model. This difference was statistically significant as per the binomial test. The comparison confidence, according to the LLM, was *high* for 739 instances (99.9%).

### Manual Evaluation

A total of 111 question–answer pairs were evaluated by ZRR and CPR. Detailed statistics comparing the performance of both systems are presented in **Table 4**.

**Table 4:**
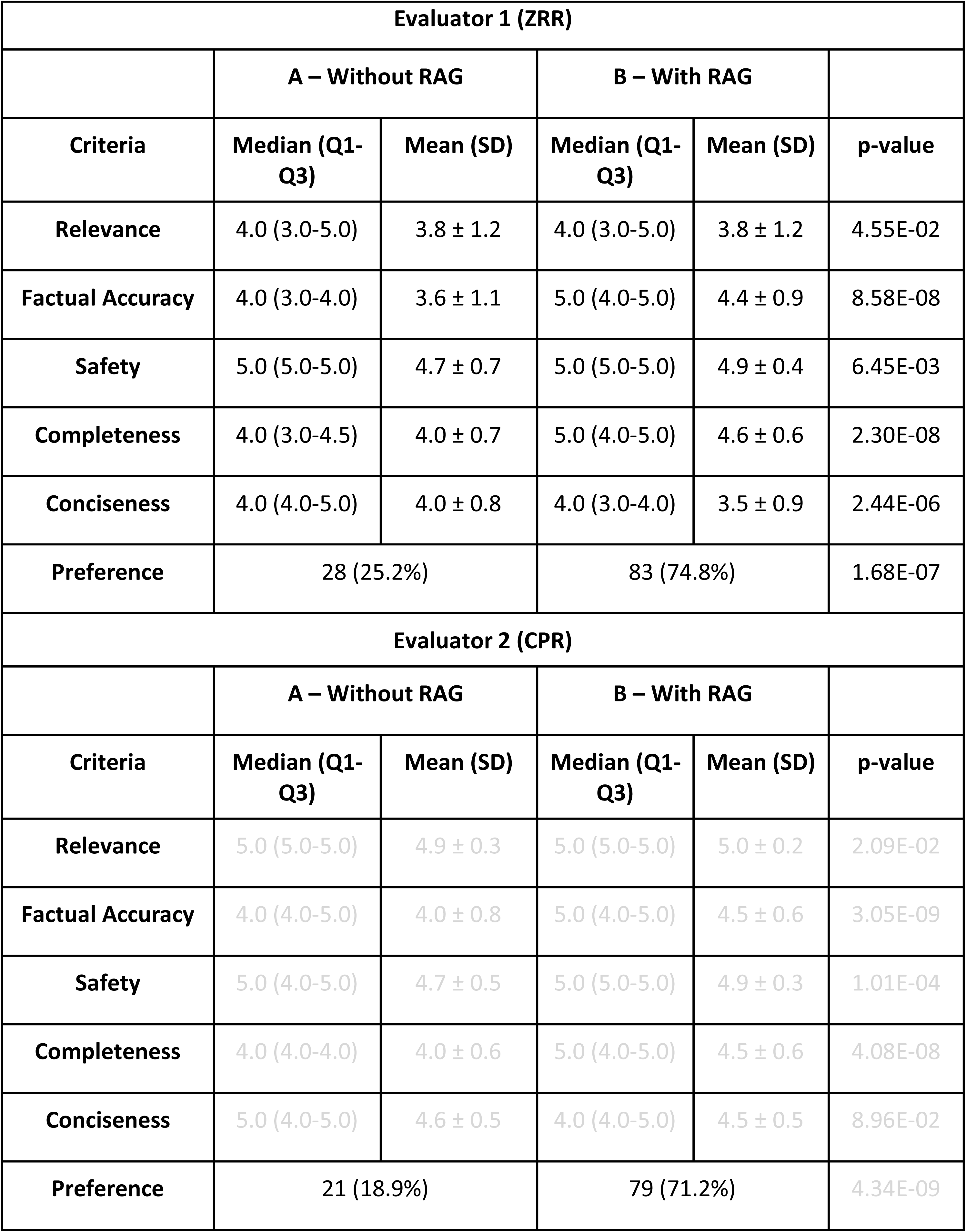
Manual evaluation results.

| Evaluator 1 (ZRR) |  |  |  |  |  |
| --- | --- | --- | --- | --- | --- |
|  | A – Without RAG |  | B – With RAG |  |  |
| Criteria | Median (Q1-Q3) | Mean (SD) | Median (Q1-Q3) | Mean (SD) | p-value |
| Relevance | 4.0 (3.0-5.0) | 3.8 ± 1.2 | 4.0 (3.0-5.0) | 3.8 ± 1.2 | 4.55E-02 |
| Factual Accuracy | 4.0 (3.0-4.0) | 3.6 ± 1.1 | 5.0 (4.0-5.0) | 4.4 ± 0.9 | 8.58E-08 |
| Safety | 5.0 (5.0-5.0) | 4.7 ± 0.7 | 5.0 (5.0-5.0) | 4.9 ± 0.4 | 6.45E-03 |
| Completeness | 4.0 (3.0-4.5) | 4.0 ± 0.7 | 5.0 (4.0-5.0) | 4.6 ± 0.6 | 2.30E-08 |
| Conciseness | 4.0 (4.0-5.0) | 4.0 ± 0.8 | 4.0 (3.0-4.0) | 3.5 ± 0.9 | 2.44E-06 |
| Preference | 28 (25.2%) |  | 83 (74.8%) |  | 1.68E-07 |
| Evaluator 2 (CPR) |  |  |  |  |  |
|  | A – Without RAG |  | B – With RAG |  |  |
| Criteria | Median (Q1-Q3) | Mean (SD) | Median (Q1-Q3) | Mean (SD) | p-value |
| Relevance | 5.0 (5.0-5.0) | 4.9 ± 0.3 | 5.0 (5.0-5.0) | 5.0 ± 0.2 | 2.09E-02 |
| Factual Accuracy | 4.0 (4.0-5.0) | 4.0 ± 0.8 | 5.0 (4.0-5.0) | 4.5 ± 0.6 | 3.05E-09 |
| Safety | 5.0 (4.0-5.0) | 4.7 ± 0.5 | 5.0 (5.0-5.0) | 4.9 ± 0.3 | 1.01E-04 |
| Completeness | 4.0 (4.0-4.0) | 4.0 ± 0.6 | 5.0 (4.0-5.0) | 4.5 ± 0.6 | 4.08E-08 |
| Conciseness | 5.0 (4.0-5.0) | 4.6 ± 0.5 | 4.0 (4.0-5.0) | 4.5 ± 0.5 | 8.96E-02 |
| Preference | 21 (18.9%) |  | 79 (71.2%) |  | 4.34E-09 |

#### Quantitative Evaluation

Both evaluators consistently rated the RAG system (B) significantly higher than the baseline (A) for factual accuracy, safety, and completeness (all p<0.001 for both reviewers). Median scores for factual accuracy and completeness were 5.0 for the RAG system versus 4.0 for the baseline. Relevance also showed statistically significant improvement for the RAG system according to both reviewers, though median scores were high for both systems. Results for Conciseness were mixed: one evaluator found the baseline significantly more concise (p<0.001), while the other found no significant difference (p=8.96×10−2).

The quantitative evaluation agreement is shown in **Table 5**. Inter-rater agreement for the quantitative scores, assessed using Kendall’s coefficient with ties and Gwet’s AC2, ranged from moderate to almost perfect. Kendall’s values (0.46–0.60) indicated moderate agreement across all criteria. Gwet’s AC2 scores suggested substantial agreement for Relevance, Completeness, and Conciseness (0.63–0.79) and substantial to almost perfect agreement for Factual Accuracy (0.76–0.83). Agreement was highest for the Safety criterion (AC2 > 0.95 for both systems).

**Table 5:** Agreement between evaluators: quantitative evaluation.

| Answer | Metric | Kendall with ties | Gwet's AC2 |
| --- | --- | --- | --- |
| A – Without RAG | Relevance | 0.51 | 0.63 |
| B – With RAG | Relevance | 0.54 | 0.66 |
| A – Without RAG | Factual accuracy | 0.59 | 0.83 |
| B – With RAG | Factual accuracy | 0.51 | 0.76 |
| A – Without RAG | Safety | 0.55 | 0.95 |
| B – With RAG | Safety | 0.46 | 0.97 |
| A – Without RAG | Completeness | 0.6 | 0.79 |
| B – With RAG | Completeness | 0.53 | 0.68 |
| A – Without RAG | Conciseness | 0.56 | 0.72 |
| B – With RAG | Conciseness | 0.6 | 0.69 |

#### Preference-Based Evaluation

In addition to scoring, the rheumatologists indicated their preferred response for each of the 111 question pairs, or marked them as comparable. Evaluator 1 favoured the RAG system in 83 instances (74.8%) compared to 28 (25.2%) for the baseline, a difference that was statistically significant (p<0.001). In contrast, Evaluator 2 rated 11 answer pairs as comparable in quality and thus did not assign a preference for these. For the remaining 100 pairs Evaluator 2 preferred the RAG system’s responses in 79 cases (71.2%) versus 21 (18.8%) for the baseline, with this preference also being statistically significant (p<0.001).

The preference-based evaluation matrix agreement is shown in **Table 6**. Regarding the preference-based evaluation, agreement between the two evaluators was observed in 76 out of 111 pairs (68.5%). This agreement was predominantly driven by instances where both evaluators preferred the RAG system’s answer (B) (65 cases). Disagreements primarily involved differing preferences between system A and B (24 pairs combined) or instances where one evaluator rated the answers as comparable.

**Table 6:** Agreement between evaluators: preference-based evaluation.

|  |  | Evaluator 1 |  |  |
| --- | --- | --- | --- | --- |
|  |  | A – Without RAG | B – With RAG | Comparable |
| Evaluator 2 | A – Without RAG | 11 | 14 | 3 |
|  | B – With RAG | 10 | 65 | 8 |

## Discussion

This study demonstrates that a RAG system integrating EULAR and ACR guidelines significantly enhances answer quality compared to a baseline LLM without retrieval augmentation. This conclusion is supported by both an automated LLM-as-a-judge evaluation and a manual assessment conducted by two independent expert rheumatologists. The RAG-based answers consistently outperformed baseline responses across key metrics in both evaluation types. Specifically, both the automatic evaluation and the manual review by rheumatologists found statistically significant improvements (p<0.001) for the RAG system in factual accuracy, safety, and completeness. Additionally, the RAG-generated responses were strongly preferred by the LLM-as-a-judge in 92.8% of comparisons and by the evaluators in 71.2%-74.8% of the comparisons, highlighting its reliability as an evidence-based clinical support tool in rheumatology.

Building on these promising performance outcomes, we introduce the first RAG system specifically designed to optimize adult rheumatology management While similar RAG approaches have been applied in other medical disciplines—such as hepatology, nephrology, and preoperative medicine—our system represents a novel, tailored application for rheumatology, with the potential to offer clinicians a specialized tool to optimize patient care. A recent scoping review identified 31 studies in which RAG was used in clinical decision support, healthcare education, and pharmacovigilance [35].

For instance, in Kresevic et al. [36], the authors integrated the “*EASL recommendations on treatment of hepatitis C: Final update of the series*” clinical guideline into a RAG framework using GPT-4 Turbo to enhance clinical decision support. They transformed non-text sources into text-based lists, standardized the guideline structure, and employed targeted prompt engineering, resulting in an accuracy of 99% compared to a baseline of 43%. Although the study focused on a single guideline, it demonstrated that structured reformatting and domain-specific prompts can markedly improve the outputs of large language models.

On the other hand, Miao et al. [37], showcased the integration of GPT-4 with the “*KDIGO 2023 Clinical Practice Guideline for the Evaluation and Management of Chronic Kidney Disease*” guideline to address specialized nephrology queries. They reformatted the text, developed a custom ChatGPT model, and evaluated its performance in specialized nephrology queries. Although only one guideline was included, the study underscored RAG’s ability to generate up-to-date, contextually relevant answers while reducing inaccuracies. Their findings highlight the importance of structured data integration and targeted prompt engineering to enhance factual precision in clinical settings.

A recent study in preoperative medicine developed and evaluated a GPT-4 based RAG model using perioperative guidelines to assess patient fitness for surgery and generate preoperative instructions [38]. The model outperformed human evaluators in predicting medical fitness for surgery (96.4% vs 86.6%) and showed high accuracy (93.0%) in determining if a patient should be seen by a nurse or doctor. The RAG system demonstrated high reproducibility, safety, and lower hallucination rates compared to some baseline LLMs. Although human evaluators were better at generating specific medication instructions, overall accuracy across various tasks was comparable. This study reinforces the potential of RAG frameworks to enhance clinical decision-making by applying specific guidelines accurately, ensuring consistency, and improving safety in complex medical workflows.

Steybe et al. [39] built GuideGPT a GPT-4–based chatbot whose RAG pipeline indexes 449 full-text publications cited in six international guidelines on medication-related osteonecrosis of the jaw (MRONJ). Across 30 prevention, diagnosis and treatment questions, answers from GuideGPT were graded by ten MRONJ experts and scored significantly higher than a temperature-matched, non-retrieval baseline for content relevance, scientific explanation and overall agreement with expert opinion, while language quality remained comparable. The system’s architecture—sentence-level embedding (text-embedding-ada-002), cosine-similarity retrieval of the top 20 contexts, and explicit hyperlinking to source PDFs—provided transparent, up-to-date rationale for each recommendation, directly addressing traceability and hallucination concerns. These findings mirror our own results in rheumatology and underscore that tailoring RAG pipelines with rigorously curated, domain-specific corpora can measurably improve accuracy, reliability and clinician trust across diverse medical sub-disciplines.

The clinical implications of this framework are significant. In a field as dynamic as rheumatology, where diagnostic criteria and treatment protocols continuously evolve, the ability to rapidly and accurately interpret guideline recommendations could substantially improve patient outcomes. By providing concise, evidence-based summaries, this system has the potential to reduce cognitive load, streamline decision-making, and enhance adherence to best practices. Moreover, this initiative aligns with broader healthcare digitalization efforts, leveraging AI and NLP technologies to optimize clinical workflows. When properly implemented, LLMs can serve as invaluable tools for documentation, patient communication, and complex diagnostic reasoning, although their use must be accompanied by rigorous validation to mitigate risks associated with algorithmic biases and outdated training data. Integrating LLMs with a RAG framework represents a significant step forward in applying AI to rheumatology guideline interpretation.

### Future directions

Future directions may involve extending the current RAG approach through the development of specialized AI agents capable of dynamically interacting with both patients and healthcare professionals [6]. By leveraging iterative feedback loops, these agents could refine treatment recommendations in real time, better adapt to nuanced clinical contexts, and ultimately improve patient outcomes. They can also integrate multimodal data and personalized patient information for more precise decision-making. The synergy between AI agents, advanced language models, and robust guideline frameworks are anticipated to substantially transform the practice of rheumatology.

Other approaches such as GraphRAG [40] or other RAG improvements such as reranking, or contextual compression could be explored [41]. However, we chose to test a simplified RAG architecture to maintain a more focused scope for our initial experiments and establish a clear baseline for future work.

### Limitations

This study is not without limitations. To start with, some recommendations contained supplementary data that were not integrated into the RAG system despite their potential relevance.

On the other hand, recent work underscores that LLMs can unintentionally produce “device-like” clinical decision support, thus falling under regulatory scrutiny as potential medical devices [43]. Although current disclaimers assert, they are not for formal medical guidance, real-world usage increasingly ventures into regulated territory. This situation underscores the need for updated frameworks to ensure safety, accuracy, and equitable outcomes when LLM-based tools are deployed. Future developments should focus on refining model outputs to adhere to approved indications and clarifying regulatory lines between simple advisory tools and full-fledged medical devices. Such balanced oversight can promote innovation in rheumatology guideline implementation while safeguarding patient care.

Although the standardization of clinical guidelines is not a novel concept [44], a primary limitation identified in this study was the absence of a uniform format and standardized structure within the guidelines and their accompanying tables. The variability in color codes, symbols, and layout conventions across recommendations compounded the complexity of automated processing, extraction, and interpretation. Establishing a uniform approach for guideline drafting and table notation could substantially improve data accuracy and consistency, thereby enhancing the reliability of RAG methods.

In this study, the chunking process was carried out using markdown delimiters, specifically hashtags. This approach, although straightforward to implement, may adversely impact RAG. Since RAG is highly dependent on the integrity of contextual relationships embedded in the source text, the absence of hierarchical preservation can lead to diminished retrieval fidelity and, consequently, reduced overall performance.

Besides our pipeline did not employ re-ranking methods. While re-ranking is a promising approach for retrieving more precise and contextually relevant chunks, it also adds complexity, increases computational overhead, and may slow down retrieval speeds. Consequently, in practical clinical settings—especially during real-time consultations— applying re-ranking frameworks could be less suitable.

Furthermore, a limitation concerning the validation of the evaluation question set warrants consideration when interpreting the performance results. Although the 740 questions were generated using GPT-4.5 to emulate clinician queries and subsequently reviewed for relevance (by DFN, DB), a systematic verification was not performed to confirm that an explicit answer for each question was present within the source EULAR/ACR guidelines. This approach potentially allows for the inclusion of questions whose answers are not fully contained, or perhaps absent entirely, within the provided documents. Such potentially out-of-scope questions could introduce an unquantified degree of noise into both the baseline and RAG evaluations. Consequently, the reported performance metrics might be influenced by questions lacking direct grounding in the source material, a factor that should be considered when assessing the system’s precise capabilities for guideline-specific information retrieval and generation.

During the manual evaluation, several RAG-generated answers began with source-framing phrases such as “According to the EULAR guideline …” or “As stated in the 2023 ACR recommendations …”. Although these provenance cues were helpful for transparency, they were not present in the baseline answers. Their presence may have subconsciously primed the expert evaluators to regard the RAG answer as more authoritative or trustworthy, pre-disposing them to select it even when the baseline response was equally accurate—or, in a few cases, better. Future studies should mask or standardise source attributions across comparison arms (e.g., by removing explicit citations or adding neutral provenance statements to all answers) to minimise this potential bias.

Finally, this study’s reliance on an LLM-based evaluation paradigm for assessing the RAG system’s outputs represents a notable limitation. Although this automated approach enables efficient, large-scale assessment, it lacks the nuanced clinical perspective provided by expert rheumatologists. Consequently, the findings may not fully capture the system’s practical utility or safety in clinical settings, as subtle inaccuracies or contextual misinterpretations might be overlooked. Future studies should incorporate human expert reviews to validate these results and ensure clinical appropriateness.

### Strengths

This study has several noteworthy strengths. First, it is, to our knowledge, the first to consolidate such an extensive set of rheumatology-specific guidelines—74 EULAR and ACR documents—into a single RAG framework. By covering a broad spectrum of diseases and clinical scenarios, the system demonstrates strong potential for real-world applicability across diverse rheumatology practices. Second, the evaluation was robust, encompassing 740 user-centric questions and employing a structured LLM-as-a-judge paradigm alongside a manual review by rheumatologists, thus balancing large-scale automated assessment with expert clinical perspectives. Third, we make the dataset of 740 questions and their corresponding RAG-generated and baseline answers available for further scrutiny and reuse, thereby promoting transparency, reproducibility, and extended research in the field. Together, these strengths underscore the practical viability and potential clinical impact of a large-scale, guideline-driven RAG system in rheumatology.

## Conclusion

This work demonstrates the successful integration of rheumatology-specific clinical guidelines into a RAG system that could assist physicians at the point of care by providing rapid and reliable access to evidence-based recommendations. To our knowledge, it is the first initiative in rheumatology to leverage such an extensive collection of guidelines, highlighting its potential impact in advancing guideline-based practice. By consolidating multiple recommendations into a single, AI-driven platform, our study sets a foundational framework for subsequent, more robust developments in RAG-based clinical decision support systems.

## Supporting information

Supplementary Excel File Additional Questions.xlsx

Supplementary Excel File Initial List of Recommendations.xlsx

Supplementary Excel File List of Recommendations.xlsx

Supplementary Excel File Questions.xlsx

## Data Availability

The clinical guidelines used in this study are freely available. All LLM prompts are documented, and the code is available upon individual request For further details or additional information, please contact the corresponding authors. Supplementary File with Questions, Answers, and the Evaluation contains the data used to evaluate the RAG system performance.

## Acknowledgements

Not applicable

## Group authorship list

Not applicable

## Author approval

All authors have seen and approved the manuscript.

## Preprint

A previous draft of this work was published as a preprint and can be found at: García, A. M., Benavent, D., Barbancho, B. M., & Núñez, D. F. (2025). Optimizing the Clinical Application of Rheumatology Guidelines Using Large Language Models: A Retrieval-Augmented Generation Framework Integrating ACR and EULAR Recommendations. medRxiv, 2025-04.

## Ethics statement

This study did not require ethics approval, as it did not involve patient data, or any information governed by the GDPR. All data employed in this research are publicly available, ensuring that no sensitive personal information was used. Moreover, the proposed system is intended solely as a support tool for rheumatologists, with final clinical decisions remaining the exclusive responsibility of qualified physicians. Therefore, no formal ethical committee approval was required for this study as only publicly available, non-patient-specific guideline documents were utilized.

## Supplementary Material

The accompanying materials for this publication are provided below.

- **Supplementary Text.** Text with technical information and further experiments conducted in this study
- **Supplementary Excel File Initial List of Recommendations.** Preliminary recommendations considered for inclusion before reaching agreement among rheumatologists.
- **Supplementary Excel File List of Recommendations.** Bibliographic details of the recommendations used in the development of the RAG (Retrieval-Augmented Generation) system.
- **Supplementary Excel File Questions.** Questions generated by ChatGPT-4.5 to evaluate the RAG architecture, with 10 questions per recommendation.
- **Supplementary Excel File Additional Questions.** Questions generated by ChatGPT-4.5 to evaluate the RAG architecture, with 30 additional questions, not analysed, per recommendation.
- **Supplementary File with Questions, Answers, and the Evaluation.** Questions used to test the RAG systems, answers with and without RAG, and evaluation files.

## Supplementary Text

### Corpus descriptive statistics

The table below presents the metrics for each chunk and clinical guideline (across all chunks). Note that this count also includes the descriptions of images generated by *o1* model.

| Metric | Value expressed as median (Q1-Q3) |
| --- | --- |
| Median number of chunks | 122 (84 - 152) |
| Median token count per chunk | 40 (33 - 48) |
| Median word count per chunk | 27.5 (20.2 - 32.5) |
| Median token count per clinical guideline | 9,621 (7,374–12,645) |
| Median word count per clinical guideline | 6,566 (5,283–8,664) |

### Questions to evaluate the baseline and the RAG answers

Initially, ten questions per clinical guideline were created with *ChatGPT 4.5*. However, 30 additional, not reviewed, questions were also created as backup. These questions are shared in **Supplementary Excel File Additional Questions.**

### Alternative experiments design

We systematically tested different combinations of embedding models, LLMs for the LLM-as-a-judge evaluation paradigm, and output lengths to assess their overall impact on the evaluation process. We focused on three key components:

- The embedding model used to generate semantic representations of the text (*voyage-3/text-embedding-3-large).* The *voyage-3* embedding model is characterized by a dimensionality of 1024 and a context capacity of 32k tokens, whereas the *text-embedding-3-large* model generates embeddings with up to 3072 dimensions and accommodates a context length of 8k tokens.
- The LLM employed as the judge to evaluate the quality of the generated answers (*o3-mini/Gemini 2.0 Flash).* The *o3-mini* model incorporates reasoning capabilities, in contrast to the *Gemini 2.0 Flash*
- Whether the LLM’s response should be constrained to a maximum of seven sentences or allowed to be unconstrained (*Max 7 sentences/no limit)*

Other alternative approaches were considered such as using a different model rather than *o3-mini* to generate the answers (e.g., *o1*/*Gemini 2.0 Flash)*, varying the score similarity threshold, adjusting the maximum number of pieces of context retrieved, applying re-ranking methods and experimenting with other chunking techniques. However, this experimentation was beyond the scope of this research.

The different combinations tested are shown in **Supplementary Table 2.** In all these experiments both answers (i.e., RAG-assisted and baseline) were generated using the *o3-mini* model. It should be noted that employing the same model for both generation and evaluation is generally discouraged due to the potential for biased assessment [45]. The results of the experiments can be seen in **Supplementary Table 3.**

### Alternative experiments results

Overall, the previous eight experiments underscore that retrieval-augmented generation (RAG) consistently outperforms the baseline across most dimensions particularly when the response length was limited to seven sentences (i.e., experiments 1–4). In this scenario, the RAG-assisted system demonstrated marked superiority, achieving statistically significant preference ratings and showing concurrent improvements across all evaluated metrics including relevance, factual accuracy, safety, completeness, and conciseness compared to the baseline. Both *o3-mini* and *Gemini 2.0 Flash* confirmed these improvements.

Conversely, under unconstrained conditions (i.e., experiments 5–8), while RAG still achieves high ratings in individual criteria such as factual accuracy and safety, its overall preference score drops to 35–50%. This suggests that, in the absence of a strict sentence limit, LLM evaluators may favour more expansive answers despite minor deficits in precision or reliability. Nevertheless, these findings demonstrate the clear value of incorporating retrieval-based strategies to enhance response quality, reinforcing the importance of considering both content constraints and user expectations when optimizing generative models in applied settings. Switching the embedding model from *voyage-3* to *text-embedding-3-large* had negligible impact on outcome distributions.

### Costs

The implementation and deployment of this RAG system incurred several usage-based costs. Three distinct types of costs were incurred:

1. Embedding costs: costs related to the embedding process. The Voyage API generated expenses proportional to the number of text chunks processed for semantic indexing. At the time of this work, the cost for the *voyage-3* embedding model was $0.06 per million tokens, with the first 200 million tokens provided free of charge.
2. LLM calls costs: calls to the LLM—both for producing an answer to clinician queries and for automatic evaluation of those answers—also accrued fees. At the time of this work, the costs for the different models were:

*a. o3-mini:* Input: $1.10 per million tokens. Output: $4.40 per million tokens
*b. Gemini 2.0 Flash:* $0.10 per million tokens. Output: $0.40 per million tokens
3. Observability costs: The *LangSmith* API introduced further costs by enabling observability and logging, generating expenses per registered trace. Currently, the platform offers 5k free traces per month, after that the cost varies from $0.5 per 1k base traces to $4.5 per 1k extended traces.

While each expense is relatively modest at small scales, these costs can accumulate with increased data volume, number of queries, and frequency of evaluation, underscoring the need to balance practical benefits with economic feasibility.

Other cost-effective alternatives exist to mitigate these expenses. For instance, local models can be deployed instead of relying on expensive API calls, which may reduce costs significantly by leveraging in-house hardware and infrastructure. Additionally, open-source solutions for observability can replace proprietary logging platforms, thereby lowering operational expenses.

### Models used

Several distinct Large Language Models (LLMs) were employed at various stages of this study to leverage their specific capabilities. Initially, *ChatGPT o1*, an advanced reasoning model, was used to generate textual descriptions for figures contained within the clinical guideline documents, ensuring this visual information was captured. Subsequently, *ChatGPT 4.5* was utilized to automatically generate the initial set of 10 evaluation questions per guideline, simulating potential user queries. For the core task of answer generation, *ChatGPT o3-mini* (referred to as LLM1) produced responses for both the Retrieval-Augmented Generation (RAG) system (incorporating retrieved context) and the baseline system (relying solely on its internal knowledge). Finally, for the automated evaluation phase, *Gemini 2.0 Flash* (referred to as LLM2) served as the ‘LLM-as-a-judge’, quantitatively scoring the quality of answers from both systems based on predefined criteria and determining preference between the RAG and baseline responses.

## Supplementary Tables

**Supplementary Table 1:** List of parsing tools/frameworks initially considered for ingesting the documents.

| Tools |  |
| --- | --- |
| <u>Azure Document Intelligence</u> | <u>MinerU</u> |
| <u>AWS Textract</u> | <u>OpenParse</u> |
| <u>Docling</u> | <u>PaddleOCR</u> |
| <u>Extractous</u> | <u>PDFPlumber</u> |
| <u>Google Document AI</u> | <u>Peslac</u> |
| <u>Graphlit</u> | <u>Preprocess</u> |
| <u>GROBID</u> | <u>PyMuPDF4llm</u> |
| <u>LlamaParse</u> | <u>RAGFlow</u> |
| <u>LLMSherpa</u> | <u>Reducto</u> |
| <u>LLMWhisperer</u> | <u>Unstructured.io</u> |
| <u>Marker</u> |  |

**Supplementary Table 2:** Additional Experiments Conducted. Responses for all experiments were generated using o3-mini.

| Experiment | Embedding model | LLM-as-a-judge | Output constraint |
| --- | --- | --- | --- |
| 1 | voyage-3 | o3-mini | 7 sentences |
| 2 | voyage-3 | Gemini 2.0 Flash | 7 sentences |
| 3 | text-embedding-3-large | o3-mini | 7 sentences |
| 4 | text-embedding-3-large | Gemini 2.0 Flash | 7 sentences |
| 5 | voyage-3 | o3-mini | No limit |
| 6 | voyage-3 | Gemini 2.0 Flash | No limit |
| 7 | text-embedding-3-large | o3-mini | No limit |
| 8 | text-embedding-3-large | Gemini 2.0 Flash | No limit |

**Supplementary Table 3:** Results of the different experiments expressed as medians (Q1–Q3) and means (SD).

| Criteria | A – Without RAG |  | B – With RAG |  | p-value |
| --- | --- | --- | --- | --- | --- |
| Experiment 1: voyage-3 / o3-mini / 7 sentences |  |  |  |  |  |
|  | Median (Q1-Q3) | Mean (SD) | Median (Q1-Q3) | Mean (SD) |  |
| Relevance | 5.0 (4.0-5.0) | 4.5 ± 0.6 | 5.0 (5.0-5.0) | 4.9 ± 0.3 | 3.30E-51 |
| Factual Accuracy | 4.0 (3.0-4.0) | 3.8 ± 1.0 | 5.0 (5.0-5.0) | 4.8 ± 0.5 | 1.18E-79 |
| Safety | 5.0 (4.0-5.0) | 4.5 ± 0.8 | 5.0 (5.0-5.0) | 5.0 ± 0.3 | 3.39E-40 |
| Completeness | 3.0 (3.0-4.0) | 3.4 ± 0.8 | 5.0 (4.0-5.0) | 4.4 ± 0.7 | 1.71E-81 |
| Conciseness | 5.0 (5.0-5.0) | 4.8 ± 0.4 | 5.0 (5.0-5.0) | 4.9 ± 0.2 | 1.70E-17 |
| Preference | 69 (10.5%) |  | 588 (89.5%) |  | 1.41E-103 |
| Experiment 2: voyage-3 / Gemini 2.0 Flash / 7 sentences |  |  |  |  |  |
| Relevance | 5.0 (5.0-5.0) | 4.8 ± 0.5 | 5.0 (5.0-5.0) | 5.0 ± 0.3 | 2.01E-23 |
| Factual Accuracy | 5.0 (4.0-5.0) | 4.4 ± 0.7 | 5.0 (5.0-5.0) | 4.9 ± 0.3 | 1.68E-57 |
| Safety | 5.0 (4.0-5.0) | 4.7 ± 0.6 | 5.0 (5.0-5.0) | 5.0 ± 0.2 | 4.80E-41 |
| Completeness | 4.0 (4.0-4.0) | 4.0 ± 0.7 | 4.0 (4.0-5.0) | 4.3 ± 0.6 | 1.05E-25 |
| Conciseness | 5.0 (5.0-5.0) | 4.8 ± 0.4 | 5.0 (5.0-5.0) | 4.9 ± 0.3 | 7.52E-11 |
| Preference | 44 (7.2%) |  | 570 (92.8%) |  | 1.18E-117 |
| Experiment 3: text-embedding-3-large / o3-mini / 7 sentences |  |  |  |  |  |
| Relevance | 5.0 (4.0-5.0) | 4.5 ± 0.6 | 5.0 (5.0-5.0) | 4.9 ± 0.3 | 9.63E-57 |
| Factual Accuracy | 4.0 (3.0-4.0) | 3.8 ± 1.0 | 5.0 (5.0-5.0) | 4.8 ± 0.5 | 3.14E-83 |
| Safety | 5.0 (4.0-5.0) | 4.5 ± 0.8 | 5.0 (5.0-5.0) | 5.0 ± 0.2 | 3.53E-45 |
| Completeness | 3.0 (3.0-4.0) | 3.4 ± 0.8 | 5.0 (4.0-5.0) | 4.5 ± 0.7 | 4.82E-84 |
| Conciseness | 5.0 (5.0-5.0) | 4.8 ± 0.4 | 5.0 (5.0-5.0) | 5.0 ± 0.2 | 3.92E-21 |
| Preference | 47 (7.25%) |  | 601 (92.74%) |  | 1.81E-123 |
| Experiment 4: text-embedding-3-large / Gemini 2.0 Flash / 7 sentences |  |  |  |  |  |
| Relevance | 5.0 (5.0-5.0) | 4.8 ± 0.4 | 5.0 (5.0-5.0) | 5.0 ± 0.1 | 1.96E-28 |
| Factual Accuracy | 5.0 (4.0-5.0) | 4.4 ± 0.7 | 5.0 (5.0-5.0) | 4.9 ± 0.3 | 4.91E-59 |
| Safety | 5.0 (4.0-5.0) | 4.7 ± 0.6 | 5.0 (5.0-5.0) | 5.0 ± 0.2 | 9.76E-41 |
| Completeness | 4.0 (4.0-4.0) | 4.0 ± 0.7 | 4.0 (4.0-5.0) | 4.3 ± 0.6 | 1.02E-26 |
| Conciseness | 5.0 (5.0-5.0) | 4.8 ± 0.4 | 5.0 (5.0-5.0) | 4.9 ± 0.3 | 1.01E-13 |
| Preference | 43 (6.92%) |  | 578 (93.07%) |  | 1.17E-120 |
| Experiment 5: voyage-3 / o3-mini / no limit |  |  |  |  |  |
| Relevance | 5.0 (5.0-5.0) | 4.8 ± 0.4 | 5.0 (5.0-5.0) | 4.9 ± 0.4 | 2.84E-05 |
| Factual Accuracy | 4.0 (4.0-5.0) | 4.2 ± 0.9 | 5.0 (5.0-5.0) | 4.8 ± 0.5 | 2.19E-42 |
| Safety | 5.0 (5.0-5.0) | 4.7 ± 0.7 | 5.0 (5.0-5.0) | 5.0 ± 0.3 | 2.65E-21 |
| Completeness | 4.0 (4.0-5.0) | 4.2 ± 0.8 | 4.0 (3.0-5.0) | 4.0 ± 0.8 | 6.08E-05 |
| Conciseness | 5.0 (4.0-5.0) | 4.5 ± 0.5 | 5.0 (5.0-5.0) | 5.0 ± 0.2 | 1.58E-68 |
| Preference | 322 (50.0%) |  | 322 (50.0%) |  | 1 |
| Experiment 6: voyage-3 / Gemini 2.0 Flash / no limit |  |  |  |  |  |
| Relevance | 5.0 (5.0-5.0) | 5.0 ± 0.1 | 5.0 (5.0-5.0) | 5.0 ± 0.2 | 5.04E-01 |
| Factual Accuracy | 5.0 (5.0-5.0) | 4.7 ± 0.5 | 5.0 (5.0-5.0) | 4.9 ± 0.3 | 8.13E-23 |
| Safety | 5.0 (5.0-5.0) | 4.9 ± 0.4 | 5.0 (5.0-5.0) | 5.0 ± 0.2 | 4.83E-17 |
| Completeness | 5.0 (4.0-5.0) | 4.7 ± 0.5 | 4.0 (4.0-5.0) | 4.0 ± 0.7 | 1.77E-64 |
| Conciseness | 5.0 (4.0-5.0) | 4.5 ± 0.5 | 5.0 (5.0-5.0) | 4.9 ± 0.2 | 3.19E-61 |
| Preference | 389 (65.2%) |  | 208 (34.8%) |  | 1.14E-13 |
| Experiment 7: text-embedding-3-large / o3-mini / no limit |  |  |  |  |  |
| Relevance | 5.0 (5.0-5.0) | 4.8 ± 0.4 | 5.0 (5.0-5.0) | 4.9 ± 0.4 | 2.39E-02 |
| Factual Accuracy | 4.0 (4.0-5.0) | 4.2 ± 0.9 | 5.0 (5.0-5.0) | 4.8 ± 0.5 | 1.45E-37 |
| Safety | 5.0 (5.0-5.0) | 4.7 ± 0.7 | 5.0 (5.0-5.0) | 5.0 ± 0.2 | 1.39E-17 |
| Completeness | 4.0 (4.0-5.0) | 4.2 ± 0.8 | 4.0 (3.0-5.0) | 3.9 ± 0.8 | 1.83E-10 |
| Conciseness | 4.0 (4.0-5.0) | 4.3 ± 0.5 | 5.0 (5.0-5.0) | 5.0 ± 0.1 | 5.10E-102 |
| Preference | 336 (50.8%) |  | 326 (49.2%) |  | 7.27E-01 |
| Experiment 8: text-embedding-3-large / Gemini 2.0 Flash / no limit |  |  |  |  |  |
| Relevance | 5.0 (5.0-5.0) | 5.0 ± 0.1 | 5.0 (5.0-5.0) | 5.0 ± 0.2 | 7.97E-01 |
| Factual Accuracy | 5.0 (5.0-5.0) | 4.8 ± 0.6 | 5.0 (5.0-5.0) | 5.0 ± 0.3 | 5.10E-20 |
| Safety | 5.0 (5.0-5.0) | 4.8 ± 0.5 | 5.0 (5.0-5.0) | 5.0 ± 0.2 | 3.77E-16 |
| Completeness | 5.0 (4.0-5.0) | 4.7 ± 0.5 | 4.0 (3.0-4.0) | 3.7 ± 0.7 | 8.23E-92 |
| Conciseness | 4.0 (4.0-4.0) | 4.0 ± 0.6 | 5.0 (5.0-5.0) | 4.9 ± 0.3 | 1.73E-108 |
| Preference | 459 (65.0%) |  | 247 (35.0%) |  | 1.23E-15 |

